# From claims to care: Machine learning algorithm to classify urinary tract infection cases using Swiss health insurance data

**DOI:** 10.1101/2025.09.29.25336862

**Authors:** Soheila Aghlmandi, Sina Shafiezadeh, Carola Huber, Pascal Godet, Heiner C. Bucher, Julia Bielicki

**Author notes:** Correspondence: Dr. Soheila Aghlmandi, PhD, Paediatric Research Center, University Children’s Hospital Basel (UKBB), Basel, Switzerland.

## Abstract

**Objectives:** To evaluate whether machine learning (ML) applied to comprehensive claims data *without diagnostic codes* can distinguish a high proportion of antibiotic treatment episodes as urinary tract infection (UTI) or non-UTI cases. Such approaches may be valuable for antimicrobial stewardship when diagnosis-linked datasets are unavailable.

**Methods:** Outpatient antibiotic prescription claims from three major Swiss insurers (2017–2020; ∼40% of the Swiss population) were analyzed. Based on clinical input, specific constellations of claims codes (e.g. positive urine culture plus typical antibiotic) were a priori assigned as indicating UTI episodes, providing the reference classification. Predictors included sex, age group, comorbidity, and diagnostic tests ordered during the episode. Four ML classifiers were tested; performance and interpretability were evaluated, with XGBoost prioritized.

**Results:** After cleaning and balancing, 38,982 records (19,491 UTI; 19,491 non-UTI) were included. XGBoost achieved an AUC of 0.94, accuracy of 87.6%, sensitivity of 79.2%, and specificity of 96.1%. Misclassification was asymmetric: 11% of non-UTI cases were labeled UTI, while 2% of UTI cases were misclassified as non-UTI. Diagnostics ordered were the strongest predictors, followed by female sex and older age.

**Conclusions:** Even in the absence of diagnosis codes, ML applied to claims data can reliably identify UTI-related prescriptions. This supports the feasibility of claims-based surveillance tools for stewardship, while in parallel highlighting the need for scalable, low-burden approaches to improve direct diagnostic coding in routine data.

## Introduction

Urinary tract infections (UTIs) are among the most common bacterial infections globally, and a leading reason for outpatient antibiotic prescribing (1, 2). In Switzerland, antibiotics prescribed for suspected UTIs rank among the most frequently dispensed agents in outpatient practice, reflecting their central role in primary care (3). While treatment is often appropriate, prolonged, unnecessary or poorly targeted antibiotic treatment remains frequent and could contribute to antimicrobial resistance, a major public health concern (2, 3). Surveillance of prescribing quality is therefore a central component of stewardship programs.

Robust surveillance usually requires diagnostic information, but this is not always available at scale. In Switzerland, for example, health insurance claims cover large segments of the population and capture all reimbursed prescribing yet lack standardized diagnosis codes. This limits their direct use for assessing appropriateness of antibiotic treatment.

Machine learning (ML) offers new opportunities to bridge this gap. By exploiting complex associations between patient demographics, comorbidity and diagnostics performed, ML can identify patterns in administrative data that correlate with clinical diagnoses (4, 5). Previous applications of ML to UTI management, including antibiotic treatment, have largely relied on electronic health records or microbiological results, but few have examined claims data, which are typically standardized, representative and widely available (6, 7).

We hypothesized that even in the absence of diagnosis codes, certain constellations of claims items, such as urine culture or other diagnostics, could serve as a reference standard for UTI-related episodes. Building on this, ML models might distinguish UTI from non-UTI antibiotic prescriptions with high accuracy.

The objective of this study was therefore to evaluate whether ML applied to comprehensive Swiss claims data *without diagnostic codes* can classify antibiotic treatment episodes as UTI or non-UTI. Demonstrating this feasibility would provide a scalable approach to surveillance in contexts where diagnosis-linked datasets are unavailable, while complementing ongoing efforts to improve direct diagnostic coding.

## Methods

### Data source and study population

We analyzed pseudonymized outpatient antibiotic prescription claims from Helsana, Sanitas, and CSS, three of the largest health insurers in Switzerland, cover approximately 40% of the Swiss population, providing a representative sample of prescribing behavior. Data spanned January 2017 through December 2020 and included all outpatient antibiotic prescriptions issued by primary care physicians (8, 9). From an initial 1,456,269 antibiotic prescriptions, records with missing, inconsistent, or implausible coding were excluded. After cleaning, 108,937 prescriptions remained. To construct a dataset suitable for ML training, treatment episodes were labeled as UTI or non-UTI based on predefined claims patterns (see below). For balanced training, UTI cases were randomly down sampled to match the number of non-UTI cases yielding 38,982 prescriptions (19,491 per group).

Because diagnostic codes are not routinely available in Swiss claims data, we defined UTI-related episodes using an a priori algorithm developed with clinical input. Specific constellations of claims items (e.g. orders for and positive urine cultures) were considered indicative of UTI. Episodes without such constellations were categorized as non-UTI. Examples of code groupings used are provided in Supplementary Methods and also Figure S1. This approach allowed construction of a reference classification that reflects real-world situations where direct diagnoses are absent.

### Predictors and model development

We included sex (male/female), age group (<5 years, ≥5–≤10, >10–≤20, >20–≤45, >45–≤65, >65), comorbidity (present/absent), and claims-based laboratory orders (basic/advanced biochemical and microbiological urinary, respiratory and blood tests), all being widely available. Four ML classifiers were implemented: eXtreme Gradient Boosting (XGBoost) (10), support vector machine (SVM), k-nearest neighbors (KNN), and naïve Bayes (10-12). XGBoost was prioritized due to its scalability, ability to handle structured datasets, and established performance in health data applications. Final hyperparameter settings for XGBoost are provided in Supplementary Table S2.

### Evaluation

Data were randomly split into training (80%) and test (20%) sets. Performance was assessed with 5-fold cross-validation on the training set to reduce overfitting. We report accuracy, sensitivity, specificity, and area under the receiver operating characteristic curve (AUC). A confusion matrix was generated for the best-performing model to examine misclassification patterns. Model interpretability was evaluated using SHapley Additive exPlanations (SHAP) to quantify the contribution of each predictor (13).

## Results

### Dataset characteristics

Of the 1.46 million outpatient antibiotic prescriptions initially available, 108,937 episodes remained after cleaning. Following classification into UTI vs. non-UTI episodes based on predefined claims constellations, a balanced dataset of 38,982 prescriptions was generated for ML training and evaluation. Descriptive characteristics are summarized in Supplementary Table S1.

### Model performance

All four classifiers achieved acceptable discrimination, with AUC under the ROC curve values >0.80. XGBoost consistently outperformed alternatives, achieving an AUC of 0.94, accuracy of 87.6%, sensitivity of 79.2%, and specificity of 96.1% on the test set. SVM (AUC=0.89), KNN (AUC=0.85), and naïve Bayes (AUC=0.82) performed less well (Figure 1).

**Figure 1.**
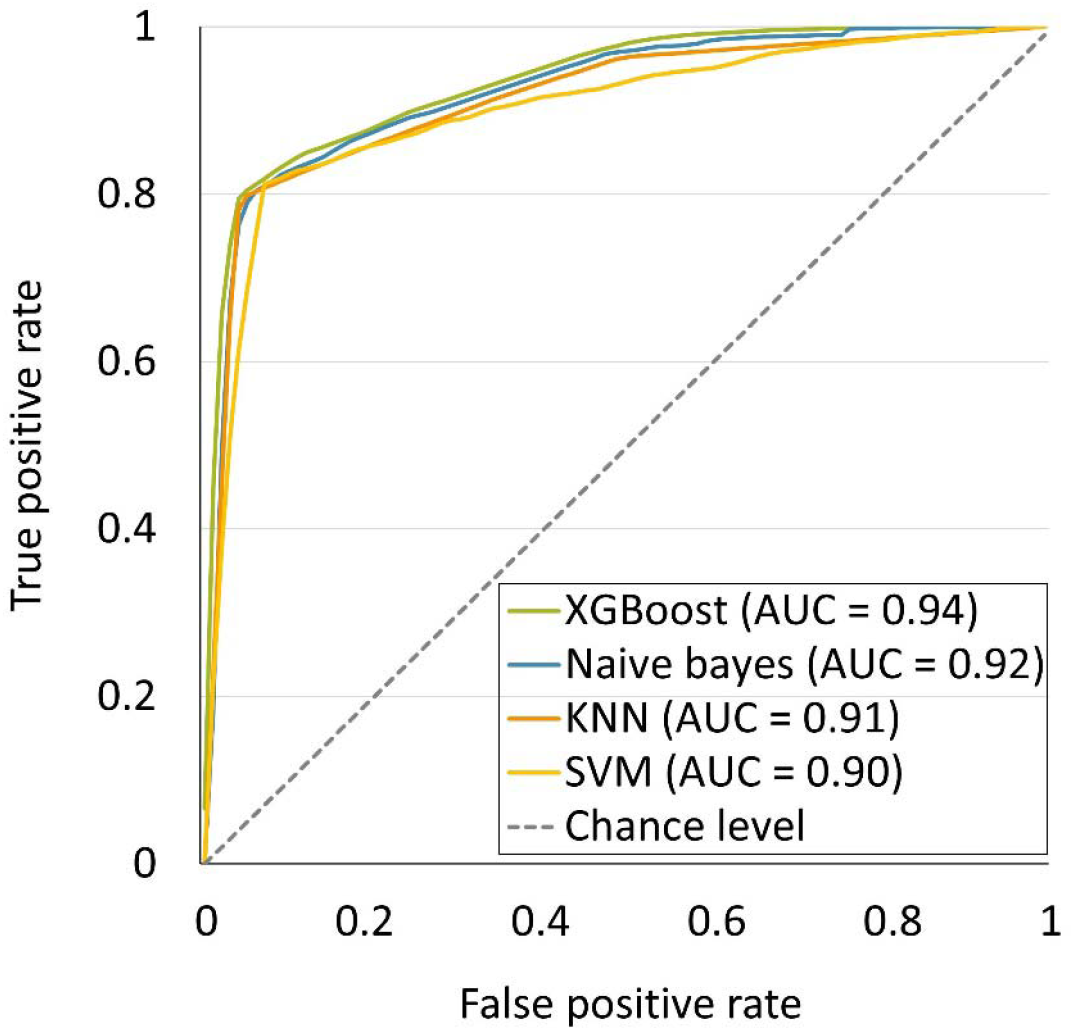
Comparison of machine learning classifiers for UTI prediction. XGBoost: Extreme Gradient Boosting, KNN: k-Nearest Neighbor, and SVM: Support Vector Machine

The confusion matrix for XGBoost revealed asymmetric misclassification. Approximately 11% of non-UTI cases were incorrectly classified as UTI, compared with only 2% of UTI cases misclassified as non-UTI (Supplementary Figure S2). This bias towards false-positive predictions suggests the model is conservative in labeling episodes as UTI, which is less problematic for surveillance than for clinical decision support.

### Feature importance

SHAP analysis demonstrated that claims-based diagnostic orders were the strongest predictor of classification, contributing most of the model explanatory power (Figure 2, and Supplementary Table S3). For example, UTI-specific tests appeared in 57.6% of UTI vs 5.2% of non-UTI cases, while general infection tests were present in 18.9% of UTI vs 75.1% of non-UTI cases (Supplementary Table S1). Female sex was the second most influential predictor, consistent with known UTI epidemiology (14). Age group also contributed significantly, with older adults expectedly accounting for the highest predictive importance (15). Interestingly, absence of comorbidity was associated with a higher likelihood of UTI classification. This likely reflects differing test use: in otherwise healthy patients, claims codes for diagnostic orders are more often directed at confirming a suspected UTI, whereas in multimorbid patients such tests may be used for broader differential purposes, making their association with UTI less specific.

**Figure 2.**
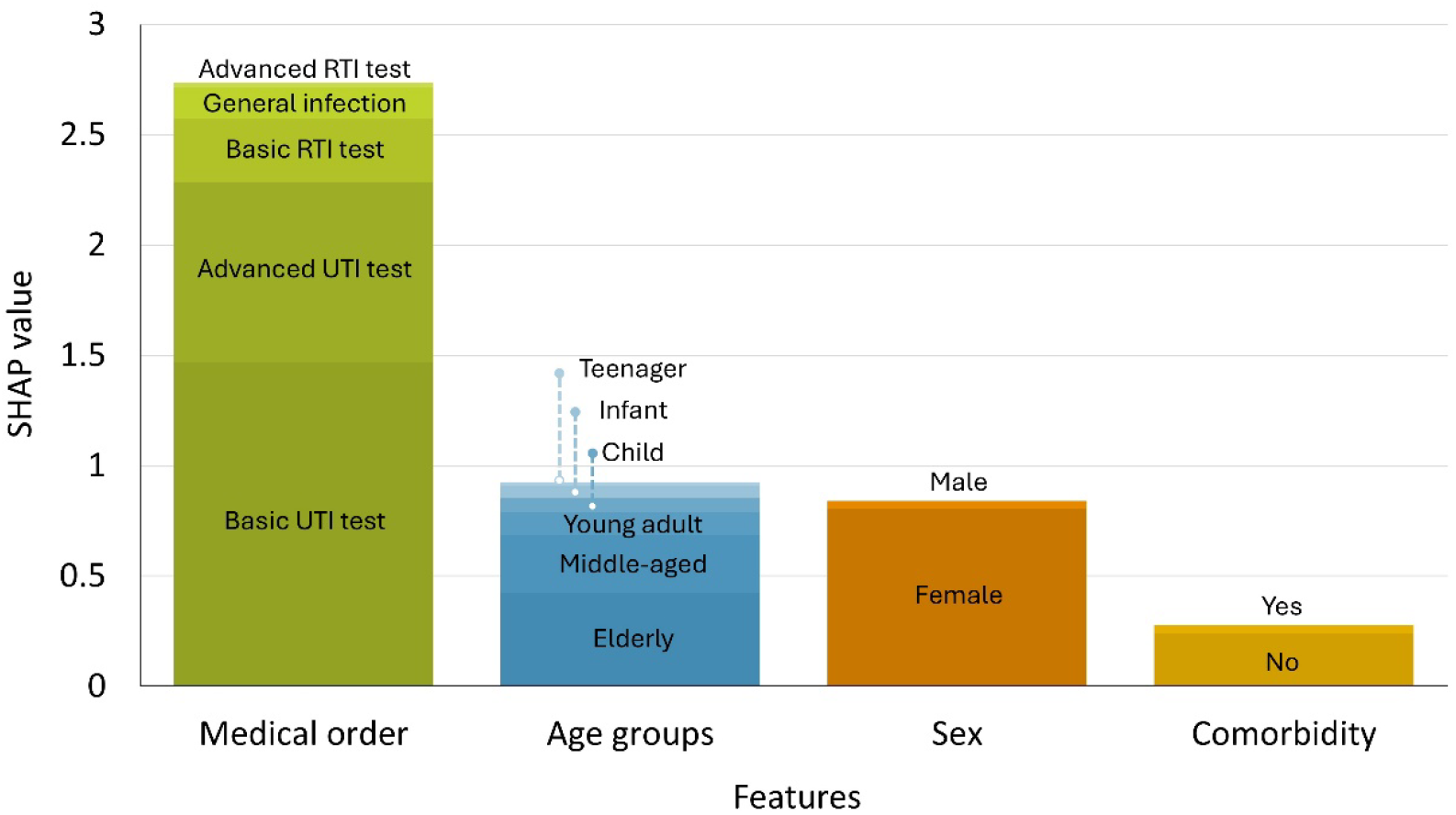
Relative importance of predictors (SHAP values) in XGBoost classification of UTI vs non-UTI episodes.

## Discussion

This proof-of-concept study demonstrates that ML applied to health insurance claims can distinguish UTI-related vs. non-UTI antibiotic treatment episodes with high accuracy, even when diagnostic codes are not available. Using predefined constellations of claims items as a reference, XGBoost achieved excellent discrimination (AUC 0.94), and feature importance analysis confirmed the face validity of key predictors: diagnostics ordered, female sex, and older age.

### Implications for stewardship

Our findings highlight the potential of claims-based ML models to support antimicrobial stewardship by enabling scalable surveillance of prescribing patterns. Large, representative datasets, such as national health insurance claims, could provide timely insights into outpatient prescribing, particularly where diagnosis-linked datasets are not available. In such contexts, ML applied to claims may be the only feasible option to approximate diagnosis-specific prescribing at population level.

At the same time, claims-based approaches should not be viewed as a substitute for improving clinical data quality. Efforts to implement low-burden diagnostic coding in routine care remain essential for precise surveillance and stewardship. Rather, claims-based ML can serve as a pragmatic interim tool, useful where diagnostic coding is incomplete, inconsistent, or unavailable, and as a complement to more granular systems once they are established.

### Strengths and limitations

Strengths of this study include use of a large, representative dataset covering nearly 40% of the Swiss population, systematic model evaluation across several classifiers and interpretability through SHAP values. Limitations include reliance on proxy definitions of UTI episodes, potential misclassification due to coding practices and lack of external validation outside Switzerland. Notably, absence of comorbidity was associated with higher likelihood of UTI classification, potentially indicative of variations in diagnostic practice across different patient strata. The observed bias towards false positives also reduces suitability or individual-level decision support or feedback, though it is less problematic in surveillance applications.

### Future work

Further research test the transferability of these models to other healthcare systems, if feasible integrate additional clinical and microbiological predictors and evaluate whether claims-based surveillance can be used to influence prescribing behavior or reduce inappropriate antibiotic use.

## Supporting information

THe document includes more information about the details of the data processing & also the methods with the relevant tables & figures

## Data Availability

All data produced in the present study are available upon reasonable request to the authors

## Transparency declaration

### Data sharing

Fully anonymized data and a data dictionary may be available with the publication of this study for purposes to be specified by applicants and with approval from the Research Committee of the programme ‘Antimicrobial Resistance’ from the Swiss National Science Foundation and with additional approval from the health insurers Helsana, CSS and Sanitas.

## Acknowledgements

We thank Helsana, Sanitas, and CSS for providing anonymized data and the Paediatric Research Center at the University Children’s Hospital Basel (UKBB) for institutional support. The results have not been presented previously at a scientific meeting or in another public context. AI-assisted editing was used to improve clarity and language; the authors remain responsible for the content.

## Access to data

The guarantor of the data is Dr. Soheila Aghlmandi, who had full access to all data used in this study.

## Contribution

Soheila Aghlmandi (SA) and Julia Bielicki (JB) conceived the study. Sina Shafiezadeh (SS) was responsible for data curation and statistical analysis. SA supervised the project and contributed to the statistical methodology. SS and SA wrote the first draft of the manuscript. JB contributed to clinical interpretation and infectious diseases expertise. JB and Heiner C. Bucher (HCB) provided methodological oversight, contributed to the epidemiological interpretation, and critically revised the manuscript. SA served as guarantor of the data.

