## Supplementary material for "From claims to care: Machine learning algorithm to classify urinary tract infection cases using Swiss health insurance data": THe document includes more information about the details of the data processing & also the methods with the relevant tables & figures

### Supplementary Methods

#### Data sources and study population

Outpatient antibiotic prescriptions were obtained from administrative claims of three major Swiss health insurers (Sanitas, CSS, Helsana). These insurers together cover approximately 40% of the Swiss population and include insured individuals across all cantons, age groups, and socioeconomic strata. The observation period was January 2017–December 2020, yielding 1,456,269 antibiotic prescriptions. This large and diverse dataset allowed for robust analysis of outpatient prescribing in Switzerland. Further details of the data are also available in Aghlmandi S. et al. prior works. (1-3)

#### Data cleaning and outcome assignment

A standardized multi-step cleaning protocol was applied to ensure internal consistency and reliability.

1. **Missing demographics:** Prescriptions without valid entries for sex or age were excluded.
2. **Implausible ages:** Records with negative values or ages >100 years were removed.
3. **Incomplete or inconsistent coding:** Prescriptions missing Anatomical Therapeutic Chemical (4) (ATC) codes , missing Swiss analysis-list codes, or containing implausible combinations of these codes were discarded.

Of note In the Swiss health system, analysis list codes are standardized billing codes on the Federal Analysis List (Analysenliste, AL) that specify laboratory tests and diagnostics reimbursed by mandatory health insurance (5).

The initial extract included 1,456,269 unique outpatient antibiotic prescriptions per patient identified with the Swiss outpatient medical tariff code system (TARMED codes) (6) LG-52 (general laboratory services) or 39.3260 (urine diagnostics). To ensure internal consistency, several exclusion steps were applied. First, prescriptions were excluded if the *analysis list code* was not among the predefined diagnostic categories relevant to infections, which removed 720,861 records. The retained codes corresponded to urine analysis, culture and sensitivity testing, and infection-related diagnostic procedures (e.g., 1245.00 urine test strip, 1372.00 urine culture, 1739.00 microbiological examination, 3469.01 rapid test, 3325.00 bacterial identification).

Second, 611,924 prescriptions were excluded because the *antibiotic ATC code* was not in the pre-specified set of systemic antibiotics relevant to urinary and respiratory tract infections. Retained classes included:

- J01CA04 (Amoxicillin)
- J01CE02 (Phenoxymethylpenicillin)
- J01D (Other beta-lactams including cephalosporins)
- J01EE01 (Cotrimoxazole)
- J01FA01 (Erythromycin)
- J01FA09 (Clarithromycin)
- J01FA10 (Azithromycin)
- J01MA02 (Ciprofloxacin)
- J01MA06 (Norfloxacin)
- J01XE01 (Nitrofurantoin)
- J01XX01 (Fosfomycin)

After these steps, 108,937 cleaned patient-level prescriptions remained.

For analytic consistency, we further restricted the dataset to prescriptions with antibiotics directly relevant for urinary tract infections or common comparators. A total of 67,510 prescriptions were excluded because their ATC codes were not part of the final classification sets: UTI-specific antibiotics (Fosfomycin J01XX01, Nitrofurantoin J01XE01) and non-UTI comparators (Azithromycin J01FA10, Clarithromycin J01FA09, Phenoxymethylpenicillin J01CE02).

To balance case groups, random down-sampling was applied to the majority class, excluding 2,445 prescriptions.

The final analytic dataset contained 38,982 balanced prescriptions (UTI and non-UTI) used for model development and evaluation. This multistep process ensured that the data retained only clinically relevant diagnostic and antibiotic codes, enabling robust and interpretable classification (Figure S1).

#### Predictors

Four main predictor groups were included:

- **Sex:** male or female.
- **Age group:** Age was categorized as <5 years, ≥5–≤10, >10–≤20, >20–≤45, >45–≤65, and >65. Age grouping enhanced clinical interpretability and ensured adequate representation within categories.
- **Comorbidity:** present or absent, derived from claims-based proxies for chronic conditions (1).

In addition, claims-based laboratory orders (basic/advanced biochemical and microbiological urinary, respiratory and blood tests) were included as diagnostic proxies (Table S1). These variables provide indirect contextual information that partially compensates for the absence of structured diagnostic coding in Swiss claims data.

#### Feature engineering and partitioning

Categorical predictors were transformed into binary variables using one-hot encoding, yielding a 19-dimensional input space. Age was included as categorical rather than continuous to facilitate interpretability and align with clinical reporting standards.

The balanced dataset was split into training (80%) and testing (20%) sets using stratified random sampling. Stratification ensured equal representation of UTI and non-UTI cases in both sets, reducing bias from unequal class distribution and improving comparability of performance across models.

#### Model development and hyperparameter tuning

We benchmarked four supervised classifiers:

1. **Extreme Gradient Boosting (XGBoost):** an efficient ensemble of gradient-boosted decision trees optimized for structured/tabular data (7).
2. **Support Vector Machine (SVM, linear kernel):** a maximum-margin classifier suited for high-dimensional spaces (8-10).
3. **k-nearest neighbors (KNN):** a non-parametric, distance-based classifier.
4. **Naïve Bayes:** a probabilistic classifier based on Bayes’ theorem, commonly applied to categorical feature spaces.

All models were implemented in Python (v3.10.12) using scikit-learn and XGBoost packages (11).

**Hyperparameter optimization** was conducted using Optuna with 100 iterations per model. Search spaces included number of estimators, maximum depth, learning rate, subsampling fraction, minimum child weight, and L1/L2 regularization. The optimal configuration for XGBoost, balancing predictive performance and generalizability, is reported in Table S2.

#### Model Evaluation

Classifier performance was assessed on the independent test set. Metrics included:

- **Accuracy:** proportion of overall correct classifications.
- **Sensitivity:** recall of UTI cases.
- **Specificity:** recall of non-UTI cases.
- **Area under the receiver operating characteristic curve (AUC):** AUC is reflecting discrimination across thresholds.

To ensure robustness, 5-fold cross-validation was performed on the training set. Mean cross-validation results were compared with test-set outcomes to evaluate stability and detect possible overfitting.

#### Interpretability

Model interpretability was evaluated using SHapley Additive exPlanations (SHAP) (11, 12). SHAP values quantify the marginal contribution of each predictor to the model’s predictions. Both global feature importance and subcategory-level contributions were computed:

- Global importance rankings identify the overall most influential predictors (Figure S2).
- Subcategory values provide detailed insight into the relative contributions of sex, age groups, comorbidity, claims-based laboratory orders, and individual antibiotic classes (Table S3).

These analyses highlight which features most strongly influenced UTI classification and which contributed only marginally, supporting transparent interpretation of model behavior.

### Supplementary Figures and Tables

#### Figures

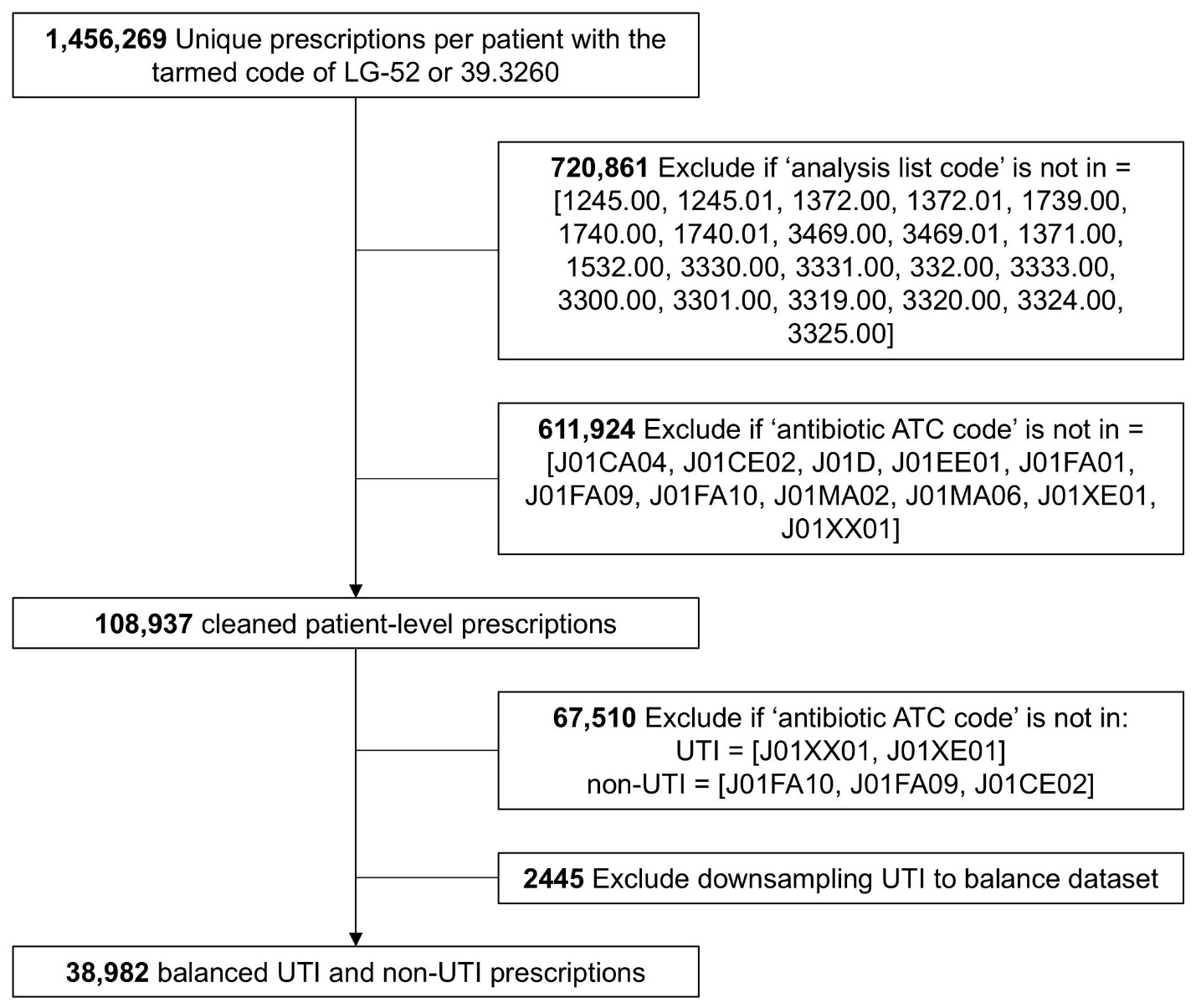

**Figure S1**. Flow diagram of data selection

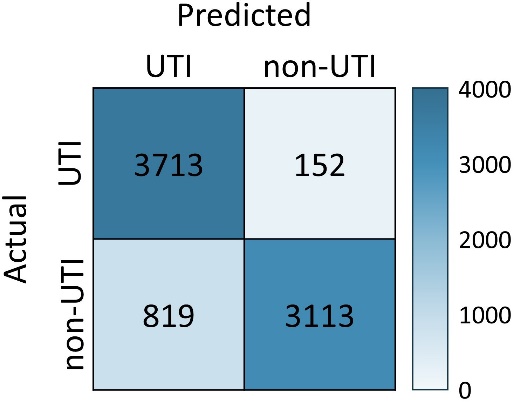

**Figure S2**. Confusion matrix showing the performance of the machine learning model in classifying UTI versus non-UTI prescriptions for XGBoost model

#### Tables

**Table S1.** The descriptive details of the claimed dataset after applying this study’s criteria to label UTI and non-UTI prescriptions.

|  | | Prescriptions, No. (%) | |
| --- | --- | --- | --- |
|  | **UTI**  N = 19,491 | | **Non-UTI**  N = 19,491 |
| Sex | |  |  |
| Female | | 16,467 (84.5%) | 11,437 (58.7%) |
| Male | | 3024 (15.5%) | 8054 (41.3%) |
| Age group (years) | |  |  |
| <5 | | 158 (0.8%) | 1190 (6.1%) |
| ≥5–≤10 | | 5818 (29.8%) | 2294 (11.8%) |
| >10–≤20 | | 0 (0.0%) | 430 (2.2%) |
| >20–≤45 | | 5335 (27.4%) | 4408 (22.6%) |
| >45–≤65 | | 1050 (5.4%) | 1672 (8.6%) |
| >65 | | 7130 (36.6%) | 9497 (48.7%) |
| Comorbidity | |  |  |
| Present | | 6334 (32.5%) | 12,062 (61.9%) |
| Absent | | 13,157 (67.5%) | 7429 (38.1%) |
| Claims-based laboratory orders | | | |
| Advanced RTI test | | 6 (0.0%) | 321 (1.6%) |
| Advanced UTI test | | 4554 (23.4%) | 271 (1.4%) |
| Basic RTI test | | 25 (0.1%) | 3247 (16.7%) |
| Basic UTI test | | 11,221 (57.6%) | 1011 (5.2%) |
| General infection | | 3685 (18.9%) | 14,641 (75.1%) |

**Table S2**. Optimized hyperparameters for XGBoost

| Search Space | Best Value |
| --- | --- |
| Number of trees(100 to 1000) | 900 |
| Maximum tree depth (3 to 10) | 9 |
| Learning rate (0.01 to 0.3) | 0.039 |
| Data subsample fraction (0.5 to 1.0) | 0.847 |
| Feature subsample fraction (0.5 to 1.0) | 0.734 |
| Minimum child weight (1 to 10) | 8 |
| Regularization alpha (1×10^−5^ to 10.0) | 5.6×10^-4^ |
| Regularization lambda (1×10^−5^ to 10.0) | 2.63×10^-5^ |

**Table S3**. SHAP values for individual features and subcategories

| **Features** | **SHAP value** |
| --- | --- |
| **Sex** | |
| Female | 0.806 |
| Male | 0.035 |
| **Age group (years)** | |
| <5 | 0.057 |
| ≥5–≤10 | 0.066 |
| >10–≤20 | 0.016 |
| >20–≤45 | 0.102 |
| >45–≤65 | 0.260 |
| >65–100 | 0.425 |
| **Comorbidity** | |
| Absent | 0.238 |
| Present | 0.039 |
| **Claims-based laboratory orders** | |
| Advanced RTI test | 0.024 |
| Advanced UTI test | 0.816 |
| Basic RTI test | 0.289 |
| Basic UTI test | 1.470 |
| General infection | 0.139 |

4.	World Health Organization. Oslo N, 2009. WHO Collaborating Centre for Drug Statistics Methodology: the Anatomical Therapeutic Chemical (ATC) and defined daily dosing (DDD) system.

12.	Lundberg SM, Lee SI. A Unified Approach to Interpreting Model Predictions. Adv Neur In. 2017;30.
